# Disinfection of SARS-CoV-2 using UVC reveals wavelength sensitivity contributes towards rapid virucidal activity

**DOI:** 10.1101/2021.06.30.21259769

**Authors:** Richard M. Mariita, Amy C. Wilson Miller, Rajul V. Randive, Lindsay G. A. McKay, Nadia Storm, Anthony Griffiths

## Abstract

SARS-CoV-2 can be disinfected using ultraviolet-C (UVC) light. For effective inactivation strategies, design and implementation, knowledge of UVC wavelength sensitivity, and disinfection rate of the relevant pathogen are required. This study aimed to determine the inactivation profile of SARS-CoV-2 using UVC irradiation with different wavelengths. Specifically, the study determined dosage, inactivation levels, and wavelength sensitivity of SARS-CoV-2. Assessment of SARS-CoV-2 (isolate USA/WA1-2020) inactivation at peak wavelengths of 259, 268, 270, 275 and 280 nm was performed using a plaque assay method. A UVC dose of 3.1 mJ/cm^2^ using 259 and 268 nm arrays yielded log reduction values (LRV) of 2.32 and 2.44, respectively. With a dose of 5 mJ/cm^2^, arrays of peak wavelengths at 259 and 268 nm obtained similar inactivation levels (LRV 2.97 and 2.80 respectively). The arrays of longer wavelength (270, 275 and 280 nm), demonstrated lower performances (≤LRV 2.0) when applying an irradiation dose of 5 mJ/cm^2^. Additional study with the 268 nm array revealed that a dose of 6.25 mJ/cm^2^ is enough to obtain a LRV of 3. These results suggest that 259 and 268 nm are the most efficient wavelengths for SARS-CoV-2 inactivation compared to longer UVC wavelengths, allowing the calculation of disinfection systems efficacy.

## Introduction

COVID-19, a pandemic disease caused by the severe acute respiratory syndrome coronavirus 2 (SARS-CoV-2), has been determined to be highly susceptible to ultraviolet light ^1, 2^. At wavelengths between 100 and 280 nm (UVC range), viral inactivation is achieved by photochemical damage to nucleic acids, which results in reduced or inhibited viral replication ^3^. Because UVC products are small and quiet, can disinfect surfaces as well as air, and are increasingly efficient, interest in their use has increased dramatically ^3, 4^, the efficacy of room sanitization by UVC has been studied ^5^ and has shown promise in the prevention of hospital-acquired infections. Recent findings have shown effective inactivation of SARS-CoV-2 by UVC irradiation at 254 nm using a commercial lamp 2. Another study, utilizing different types of culture media, has demonstrated that shorter wavelengths are more effective at inactivating SARS-CoV-2 than longer wavelengths (265 nm>280 nm>300 nm) ^6^. This has led to the general understanding that UVC irradiation, especially in the range between 200-290 nm, has great inactivation potential ^7^.

Unfortunately, the data currently available do not establish an inactivation profile of SARS-CoV2 or wavelength sensitivity. Consequently, system design and implementation requires estimating how much UVC light is required to achieve desired inactivation levels. It also prevents the use of a surrogate to SARS-CoV-2 in efficacy testing, thus requiring dangerous, expensive, and difficult tests for validation. In the present study, we describe experiments conducted to investigate the wavelength sensitivity of SARS-CoV-2 on plastic surfaces. The study investigated the inactivation rates of SARS-CoV-2 at a range of UVC wavelengths (259, 268, 270, 275 and 280 nm) with similar doses in a controlled and repeatable manner.

## Results and discussion

The study developed an irradiation apparatus (Fig. 1A) to quantitatively analyze inactivation of SARS-CoV-2 with different wavelengths. A USB4000 photospectrometer (Ocean Optics) was used to confirm the emitted radiation peak wavelengths of the UVC LED arrays at 259, 268, 270, 275 and 280 nm (Fig 1B). The UVC dose was confirmed using a X1 handheld optometer (Gigahertz-Optik) (Table 1).

**Figure 1:**
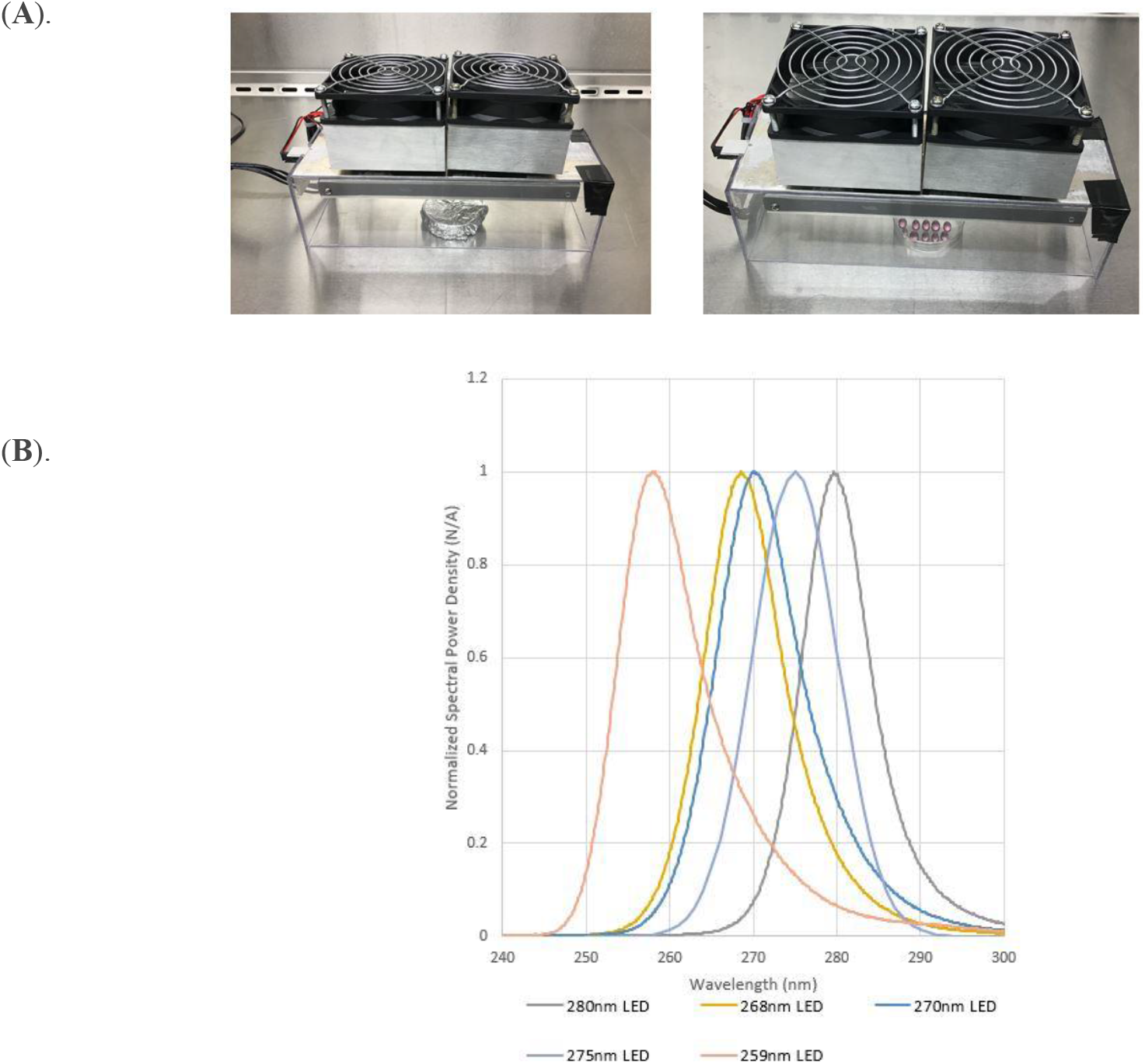
(**A**) SARS-CoV-2 array set-up for inactivation study. A pair of dishes (one to be irradiated and one control wrapped tightly in aluminum foil) were placed in the chamber of the device and UVC-irradiated for a specified number of seconds, with each irradiation time tested in triplicate. Plaque assay technique was used to test the performance of the arrays. (**B**) Confirmation of wavelength peak emissions for each array used in the study.

**Table 1:** Timepoints, UVC doses obtained at a given distance, and irradiation at different LED wavelengths revealed lower inactivation of SARS-CoV-2 at >270 nm compared to 259 and 268 nm.

| UVC wavelength (nm) | 259 |  |  | 268 |  |  | 270 |  |  | 275 |  |  | 280 |  |  |
| --- | --- | --- | --- | --- | --- | --- | --- | --- | --- | --- | --- | --- | --- | --- | --- |
| Dose (mJ/cm<sup>2</sup>) | 1.5 | 3.1 | 5 | 1.6 | 3.3 | 6 | 1.6 | 3.3 | 5 | 1.6 | 3.3 | 5 | 1.6 | 3.3 | 5 |
| Distance (cm) | 7 | 7 | 7 | 14 | 14 | 14 | 7 | 7 | 7 | 7 | 7 | 7 | 7 | 7 | 7 |
| Time (Seconds) | 7 | 15 | 23 | 3 | 6 | 11 | 4 | 8 | 13 | 4 | 8 | 12 | 4 | 8 | 12 |
| Mean control virus titer (PFU/mL) | 7500 | 5250 | 7750 | 10667 | 7333 | 9500 | 2727 | 2637 | 3237 | 3203 | 2427 | 2007 | 2480 | 2487 | 2600 |
| Mean irradiated virus titer (PFU/mL) | 217 | 25 | 8 | 383 | 27 | 15 | 377 | 45 | 31 | 912 | 83 | 73 | 1037 | 500 | 237 |
| Log reduction value (LRV) | 1.54 | 2.32 | 2.97 | 1.44 | 2.44 | 2.80 | 0.86 | 1.77 | 2.02 | 0.55 | 1.47 | 1.44 | 0.38 | 0.69 | 1.04 |

SARS-CoV-2 showed high susceptibility to UVC radiation, especially at peak emissions of 259 and 268 nm (Fig. 2). There was a strong association between wavelength and inactivation efficacy of the test arrays. Specifically, within UVC doses of 1.5 mJ/cm^2^, 3.1 mJ/cm^2^ and 5 mJ/cm^2^, there were significant differences in inactivation efficacy between arrays (*p*=0.0245, R^2^=0.8551, *p*=0.0497, R^2^=0.7723 and *p*=0.0186, R^2^=0. 8788 respectively). Inactivation performances increased with UVC exposure (dose) (Fig. 3). Additionally, because the performance of 259 and 268 nm arrays was similar, the study expanded to confirm that a LRV of 3 could be obtained by applying a dose of 6.25 mJ/cm^2^ in 5 seconds using the 268 nm array. SARS-CoV-2 was reduced to below detectable levels within 7 seconds at a UVC wavelength of 268 nm.

**Figure 2:**
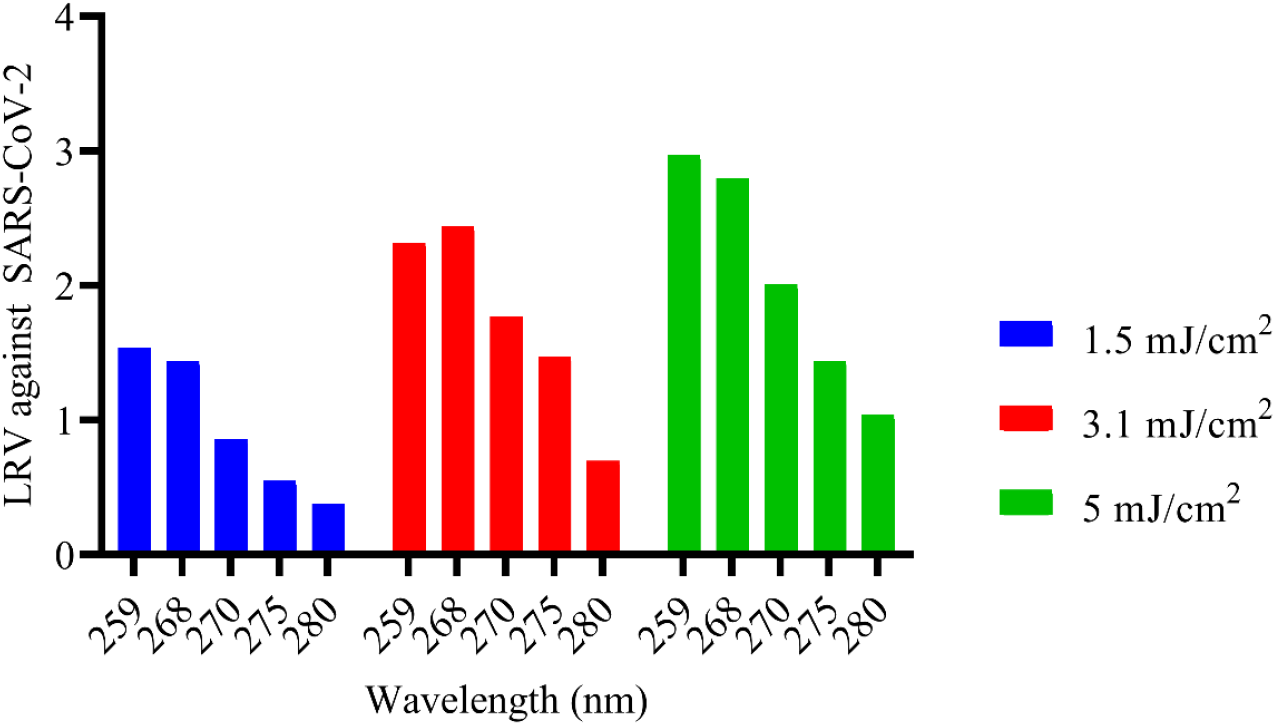
Effects of UVC LEDs with different peak emissions on SARS-CoV-2 (isolate USA/WA1-2020) inactivation. Inactivation efficacy of wavelengths was carried out at similar UVC doses. Inactivation of SARS-CoV-2 revealed wavelength sensitivity, with the 268 nm array obtaining comparable performance with the 259 nm device.

**Figure 3:**
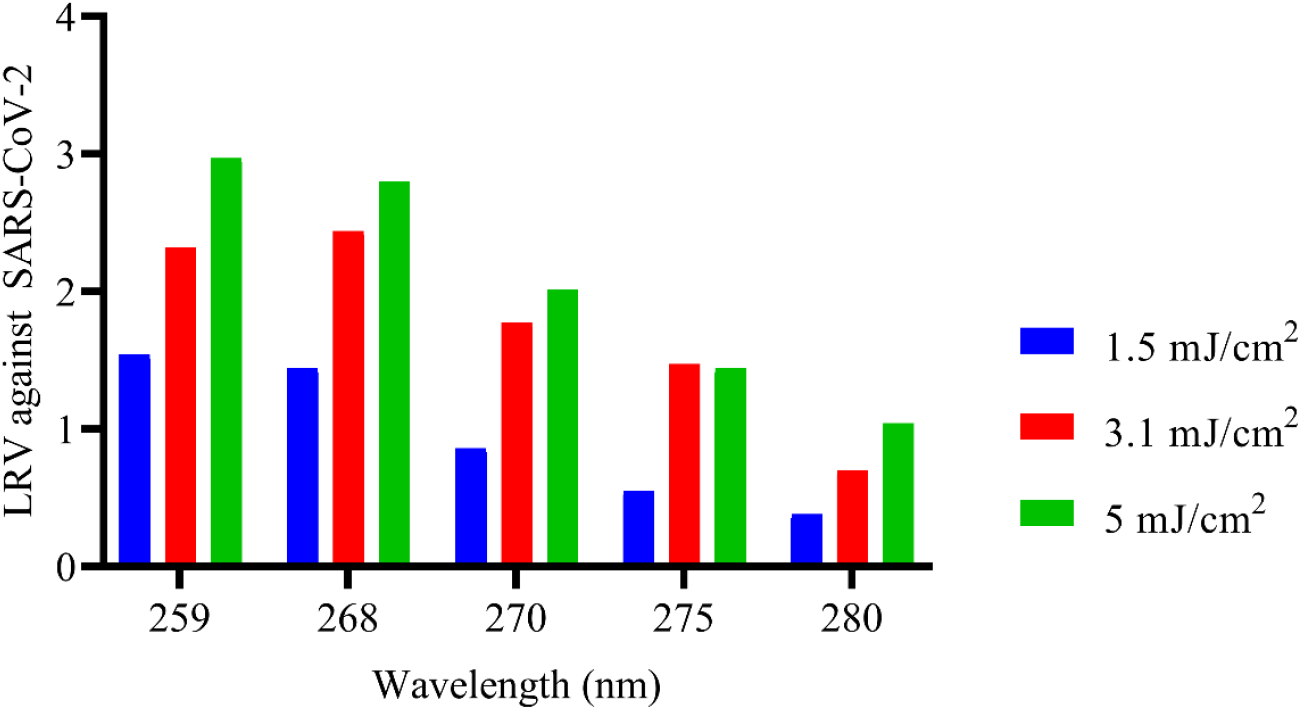
Effects of low UVC doses on SARS-CoV-2. Increases in UVC dose led to increased viral inactivation.

Findings from the current study are consistent with previous findings that UVC is very effective at inactivating SARS-CoV-2, that shorter wavelengths are more effective, and reveal the approximate required dosing ^2, 1^. These results are somewhat consistent with a study of 280 nm in which a dose of 37.5 mJ/cm^2^ was required to obtain LRV 3 against SARS-CoV-2, indicating that longer wavelengths were much less effective at inactivating the virus ^8^. For emphasis, shorter wavelengths easily achieve target inactivation while applying a smaller UVC dose ^9^. Furthermore, unlike previous wavelength sensitivity study ^10^, the current study employing SARS-CoV-2 and utilizing more wavelengths enabled a deeper assessment of their inactivation efficiency under similar conditions, leading to sensitivity determination. This confirms the cumulative benefit of UVC with time even at low UVC doses ^11^ which observed a potentially compounding benefit of UVC when used over time in long-term care facilities.

This study did not explore the efficacy of UVC in inactivating SARS-CoV-2 in particulate or aerosolized format, or on different surface types. Moreover, the experiments described in this study did not account for changes in humidity and temperature, which could potentially influence inactivation efficacy. Additional studies are required to determine the required dose for effective virus inactivation in these scenarios. Nevertheless, this study supports the design and implementation of many new UVC systems based on the inactivation profiles found for varying UVC radiation sources.

## Methods

A USB4000 photospectrometer (Ocean Optics, https://www.oceaninsight.com/) and X1 handheld optometer (Gigahertz-Optik, https://www.gigahertz-optik.com/en-us/product/x1/) were used to confirm emitted radiation peak wavelength and UVC dose of the UVC arrays, respectively. Experiments were carried out in a biosafety level 4 laboratory at the National Emerging Infectious Diseases Laboratories of Boston University (https://www.bu.edu/neidl/). Specifically, 100 μL SARS-CoV-2 (7.33 × 10^3^ PFU/mL) (USA/WA1-2020) ^12^ was plated onto the surface of 60 mm plastic coupons (tissue culture dishes) in 5 μL aliquots. The virus was then dried in the biosafety cabinet in the dark before irradiating with UVC LED arrays at a specific height, dose and time (Table 1).

A pair of tissue culture dishes (one to be irradiated and one control wrapped tightly in aluminum foil) were placed in the chamber of the device and UVC irradiated for a specified number of seconds, with each irradiation time tested in triplicate and averages used in statistical analyses. Following treatment, the virus was resuspended in 2 mL high glucose Dulbecco’s Modified Eagle Medium (DMEM) (Gibco) containing 0.04 mM phenol red, 1 × antibiotic-antimycotic (Gibco), 1 × non-essential amino acids (Gibco), 1 × GlutaMAX-I (Gibco), 1 mM sodium pyruvate (Gibco) and 2% fetal bovine serum (FBS)(Gibco). The resuspended virus was then serially diluted from 1 × 10^0^ to 1 × 10^−2.5^ using half-logarithmic dilutions before performing a semi-solid overlay plaque assay as previously described ^13^. Quantitation of plaques was carried out to determine viral titer, which was used to calculate the log reduction value (LRV) between irradiated and control samples.

*Virus titer in PFU/mL = Number of plaques /virus dilution in well × volume plated in mL* ^2^ *and* 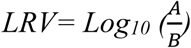

*Where A=PFU/mL for UVC controls (control wrapped tightly in aluminum foil) B =PFU/mL for UVC dosed (UVC irradiated)*

Our findings provide evidence that SARS-CoV-2 is more sensitive to UVC inactivation at shorter wavelengths (<268 nm, as opposed to >270 nm). UVC LEDs with the most effective peak emissions could serve as effective and rapid tools in the fight against SARS-CoV-2 and other pathogenic viruses. This is an important result for designing UVC-based solutions to combat infectious diseases, as engineers need to consider the effectiveness of the wavelengths used and the tolerance around rated peak wavelength.

## Data Availability

Original SARS-CoV-2 disinfection data and laboratory report is available upon reasonable request.
Spectral confirmation data is available via https://doi.org/10.6084/m9.figshare.14884743.v1

https://doi.org/10.6084/m9.figshare.14884743.v1

## Data availability

Original SARS-CoV-2 disinfection data and laboratory report is available upon reasonable request. Spectral confirmation data is available via https://doi.org/10.6084/m9.figshare.14884743.v1

## Author Contributions

R. M. M., A. C. W. M., R. V. R., N. S., L. G. A. M., A. G.: study design; N. S., L. G. A. M.: performed the experiments; R. M. M., N. S., L. G. A. M.: data analysis; R. M. M., A. C. W. M.: manuscript writing and data interpretation; R. V. R.: supply of study materials; R. M. M., A. C. W. M., R. V. R., N. S., L. G. A. M., A. G.: read, revised and approved final manuscript.

## Acknowledgements

The authors are grateful to Dr. Kevin Kahn and James Davis for proofreading the manuscript.

